# Modelling COVID-19 Vaccine Breakthrough Infections in Highly Vaccinated Israel – the effects of waning immunity and third vaccination dose

**DOI:** 10.1101/2022.01.08.22268950

**Authors:** Anyin Feng, Uri Obolski, Lewi Stone, Daihai He

**Author notes:** Equally contributed first authors (Anyin Feng, Uri Obolski). Email of all authors: AF UO LS DH. Corresponding author: Daihai He, Department of Applied Mathematics, The Hong Kong Polytechnic University, Hong Kong, China.

## Abstract

In August 2021, a major wave of the SARS-CoV-2 Delta variant erupted in the highly vaccinated population of Israel. The Delta variant has a transmission advantage over the Alpha variant, and thus replaced it in approximately two months. The outbreak led to an unexpectedly large proportion of breakthrough infections (BTI)-- a phenomenon that received worldwide attention. The BTI proportion amongst cases in the age group of 60+ years reached levels as high as ∼85% in August 2021. Most of the Israeli population, especially those 60+ age, received their second dose of the vaccination, four months before the invasion of the Delta variant. Hence, either the vaccine induced immunity dropped significantly or the Delta variant possesses immunity escaping abilities. In this work, we analyzed and model age-structured cases, vaccination coverage, and vaccine BTI data obtained from the Israeli Ministry of Health, to help understand the epidemiological factors involved in the outbreak. We propose a mathematical model which captures a multitude of factors, including age structure, the time varying vaccine efficacy, time varying transmission rate, BTIs, reduced susceptibility and infectivity of vaccinated individuals, protection duration of the vaccine induced immunity, and the vaccine distribution. We fitted our model to the cases among vaccinated and unvaccinated, for <60 and 60+ age groups, to address the aforementioned factors. We found that the transmission rate was driven by multiple factors including the invasion of Delta variant and the mitigation measures. Through a model reconstruction of the reproductive number R_0_(*t*), it was found that the peak transmission rate of the Delta variant was 1.96 times larger than the previous Alpha variant. The model estimated that the vaccine efficacy dropped significantly from >90% to ∼40% over 6 months, and that the immunity protection duration has a peaked Gamma distribution (rather than exponential). We further performed model simulations quantifying the important role of the third vaccination booster dose in reducing the levels of breakthrough infections. This allowed us to explore “what if” scenarios should the booster not have been rolled out. Application of this framework upon invasion of new pathogens, or variants of concern, can help elucidate important factors in the outbreak dynamics and highlight potential routes of action to mitigate their spread.

## Introduction

The COVID-19 pandemic caused tremendous impact globally, but would likely have led to far more damage if effective vaccines against SARS-CoV-2 had not been rapidly developed and deployed. In addition, the emergence of new SARS-CoV-2 variants having higher transmissibility and potential for immune evasion (1) posed new challenges to mitigating the pandemic through vaccination. In this respect, Israel serves as an excellent case-study of vaccine effectiveness, as it has been a frontrunner in the vaccination campaign: the first-dose vaccination coverage, which began on 19 December 2020, exceeded 60% population coverage by March 22, 2021; and the second dose vaccination (fully vaccinated), began on January 9, 2021 and exceeded 60% population coverage by July 11, 2021. The vaccine was first prioritized for the elderly, while the government approved vaccination for all adolescents 11-18 since June 2021 (2, 3), and children from 5-11 years of age since November 14, 2021. The most commonly administered vaccine in Israel is the BNT162b2 vaccine, which has high estimated efficacy (>92%) against infection and against severe outcomes from the wild-type strain (4). However, despite the high vaccine coverage and purported efficacy, a large proportion of vaccine breakthrough severe cases unexpectedly appeared in Israel during August 2021 with the new invasion of the Delta variant. Fortunately, after July 31, the government had already begun providing a third “booster” dose of the vaccine, initially targeting the elderly (5).

The complex dynamics of the pandemic in Israel are illustrated in Figure 1. The figure shows that the pandemic unfolded in four waves from March 2020 to November 2021, that were controlled by “on-and-off” lockdown and mitigation measures (which may be quantified by a “stringency index”, see SI4), strain replacement and the country’s increasing cumulative vaccination coverage. Three strains of SARS-COV-2 dominated at different times in Israel: the wild-type strain before Jan 2021, the Alpha variant (B.1.1.7) between January 2021 and June 2021, and the Delta variant (B.1.617.2) after June 2021 (6). The first two waves were caused by the wild-type strain, while the third wave coincided with the transition from the wild strain to the Alpha variant. As mentioned, the fourth wave included a large proportion of breakthrough infections (BTI), which occurred in parallel with the arrival of the Delta variant.

**Figure 1.**
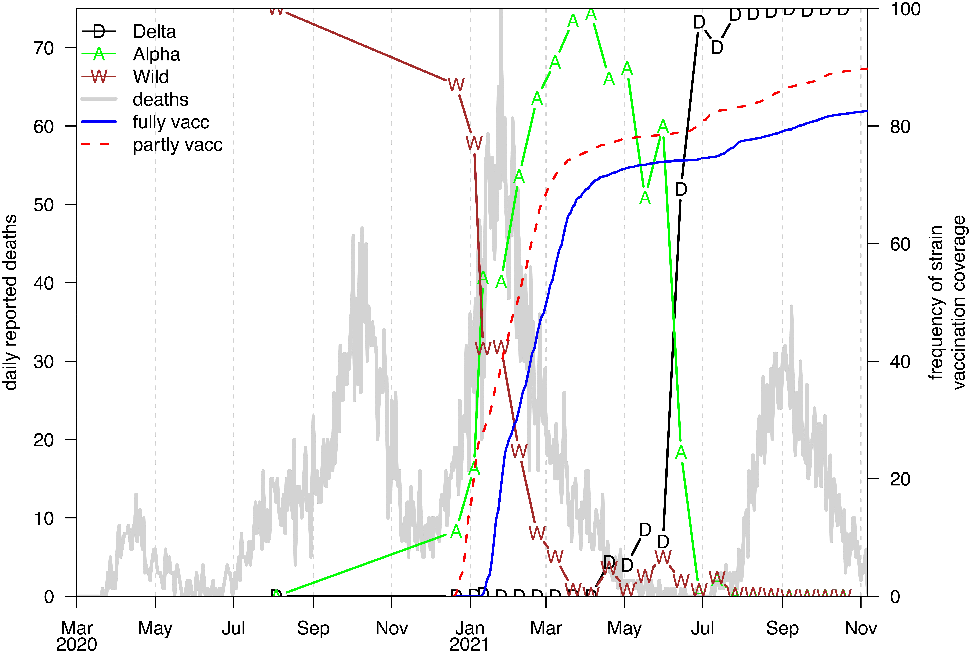
Daily reported COVID-19 deaths (grey) over pandemic period. Colored symbols show frequency of individual strains (as a percentage) of all strains sequenced (mostly biweekly) in Israel. Also plotted is the vaccination coverage (ie. proportion of the population being vaccinated) with solid blue representing fully vaccinated and red dashes for partially vaccinated (one dose). The Alpha variant replaced the wild-strain in December 2020-Febrary 2021, while the Delta replaced the Alpha variant in June-July 2021. The final wave in the graph is almost completely due to the Delta variant.

In addition to vaccination, the Israeli government issued various mitigation policies including a national lockdown from December 27, 2020 to February 5, 2021. But once the concurrent rapid vaccination campaign reduced the number of infections and deaths, the government removed most coronavirus restrictions. Nevertheless, the restrictions on gatherings were reinstated on July 29, 2021 when only people who were vaccinated or had recently tested negative were permitted to enter public places (7-9).

Several studies sought to examine the effectiveness of the BNT162b2 vaccine in Israel. Haas et al. (10) analysed national surveillance data in Israel between January 24 to April 3, 2021, and estimated an effectiveness of about 95% at 7 days or more after the second dose against infection. Goldberg et al. (11) compared the risk of infection in early July 2021, among the elderly (age 60+ years) who received the second dose of vaccine in March 2021 (four months after becoming fully vaccinated) compared to those who received the second dose vaccine in January 2021 (six months after fully becoming vaccinated). They found that the protection against infection decreased from 73% for those in the March group to 57% for those in the January group, while the efficacy against severe symptoms deteriorated from 91% to 86% in the same setting.

As mentioned above, one of the dominant factors of the fourth wave in Israel was the emergence of the Delta variant. It was quickly understood that the Delta variant had enhanced transmissibility (12) and poorer prognoses (13) than the wild type strain. Accordingly, when the Delta variant reached Israel in August 2021 the number of cases and hospitalizations increased significantly (5). As of September 13, 2021, among the 664 severe cases in Israel, 25% patients received two doses of vaccine, 65% patients were completely unvaccinated, and only 8% had received the booster shot (14).

Vaccine effectiveness against the Delta strain has been vigorously investigated since the strain’s emergence. However, the effectiveness seems to be highly dependent on, and hard to disentangle from, the time elapsed since vaccination. For example, a study of the UK population, largely within four months of vaccination at the time, estimated effectiveness of two doses of the BNT162b2 vaccine against infection with the Delta variant at 88%. On the other hand, Israel’s Ministry of Health (MoH) reports estimated between 64% and 39% effectiveness (15), when observing the population approximately 5 and 7 months after vaccination (16). This has been echoed in other studies in Israel, describing a combined effect of waning immunity and Delta strain invasion on vaccine effectiveness (11, 17).

In light of the studies above, it is unclear what are the main drivers behind the current COVID-19 wave and the observed high BTI ratio in Israel. Hence, here we present a model which takes into account the age difference in vaccination, BTI ratio, the protection duration (and distribution) of vaccine induced immunity, time varying transmission rate, and the effectiveness of the second and third dose vaccination doses.

## Materials and Methods

### Data

The following data sources, obtained from the Israel Ministry of Health (MOH) and from the World Health Organization (WHO), formed the basis of this modelling study

1. Weekly reported, PCR-confirmed cases stratified by age from March 21, 2020 until November 6, 2021 (18).
2. Daily hospitalization condition, including patients’ mean age, gender, and severity of disease from March 11, 2020 until November 6, 2021 (19).
3. Daily vaccination with three doses stratified by age, from December 20, 2020 until November 6, 2021 (20).
4. Daily diagnosed severe cases stratified by age, including patients’ status of vaccination, from July 29,2021 until November 6, 2021 (21).
5. Daily confirmed cases and deaths for all ages from March 1, 2020 until November 6, 2021 (22).
6. Proportions of the different variants of concern confirmed over time (23).

The definitions relating to vaccination used here conform to those applied by Israel’s MOH (24): individuals are considered fully vaccinated if they received 2 doses of vaccine and if 7 days had passed since the second dose; or if they had been infected with COVID-19 before they received a dose of the vaccine, and 7 days had passed since the first dose of the vaccine; partially vaccinated individuals were those who received the first dose of vaccine but not the second dose and those who have received the second dose for less than 7 days. Unvaccinated individuals are those who had not received any vaccine dose.

### Model

The model formulation used here is an elaboration of the conventional SEIRV epidemic population model where the variables *S,E,I,R* and V_(k)_ denote the number of individuals in the Susceptible, Exposed, Infectious, Recovered and Vaccinated groups. Susceptible individuals flow through the compartments in linear order: S→E→I→R. A proportion 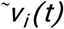 of susceptible individuals are shunted to the vaccination compartments. As explained in more detail shortly, in order to model the waning of immunity, the vaccinated class V_(k)_ is broken down into a series of five stages (*k=*1,2,3,4,5; see (25)). The model includes age-classes with *i=0,1* indicating <60 years and 60+ years, respectively. The population of Israel was approximately 9.28 million people in 2021, and the 60+ age group accounted for 16.4% of this number. The two age classes were chosen due to data availability (see datasets above) and due to interest in the elderly component of the population.

A special feature of the model is the incorporation of BTIs, which arise in those vaccinated individuals in the *V*_*(k)*_ (vaccinated) group where they at first gain medium-term temporary immunity. When this protection wanes, they join the *S*_*V*_ class of post-vaccination susceptible and become susceptible once more, and have the potential to generate BTI’s. We assume those individuals in *S*_*V*_ have partial protection (i.e., reduced susceptibility) to infection, as controlled by the parameter ε. Upon infection, they join the breakthrough exposed class, *E*_*B*_. For simplicity, we did not track BTIs after their infection, and just allowed individuals in *E*_*B*_ to join the *I* class after their latent period. Realistically however, these BTIs have reduced transmissibility and reduced severity when infected. These effects were modelled by introducing one more parameter which reduces transmissibility in BTIs (ω), which is discussed in supplementary section SI2.

The full model is given by the following ordinary differential equation system:

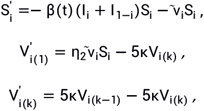

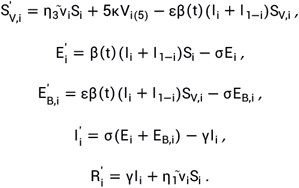

The parameters were set to reflect the most current knowledge of COVID-19 dynamics. In particular, exposed individuals move to the infectious class after a mean latent time of *1/* σ =2 days. Infected individuals remain infectious for a mean time of 1/γ= 3 days (26-28). This gives a short mean generation time of 5 days, which is the sum of the mean latent period and the mean infectious period. The model takes into consideration imperfect reporting, by assuming that only a proportion ρ_*i*_ of infected individuals are reported in each age-class i. The model infers ρ_*i*_ with the assumption that ρ*i* ≥ *0*.*4* to reflect the high medical accessibility rate in Israel.

A schematic flowchart of the model is given in Figure 2.

**Figure 2.**
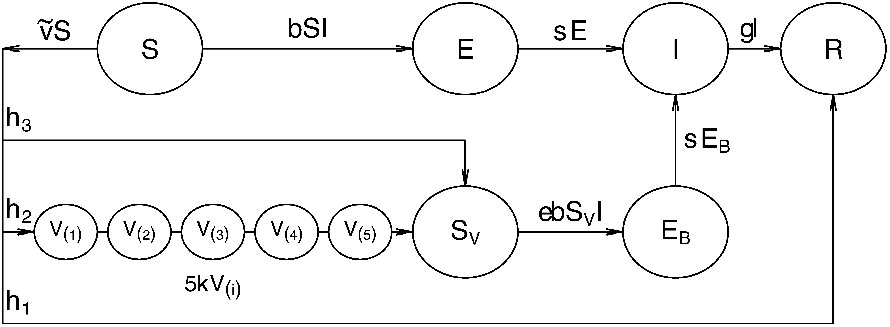
A flowchart of Susceptible-Exposed-Infective-Recovered-Vaccinated (SEIRV) model for one age group. Susceptible individuals flow through the compartments in linear order: S→E→I→R. Vaccinated susceptible individuals are of three immunity types: a proportion (η_*1*_) move directly to the Recovered class and gain long-term protection; a proportion (η_*2*_) gain medium-term temporary protection and move through a series of five V_(k)_ classes. After a period in which immunity wanes, they then move on to the *S*_*V*_ class. Individuals in *S*_*V*_ are susceptible to breakthrough infections and move to the *E*_*B*_ and *I* class; a proportion (η_*3*_) of those vaccinated have little protection and move to *S*_*V*_ class directly.

### Time varying transmission rate

Over the study period, the country’s mitigation measures, public risk perception, seasonality, and behaviors changed (as reflected in the stringency index, see SI4), and thereby continuously modified the contact rates, and thus ultimately the transmission rate. Additionally, the invasion of variants having increased transmissibility are able to modify the transmission rate in our model.

Hence, these changes need to be reflected through a time varying transmission rate β (*t)*, (following our previous work (28)).In practice, β (*t*) is obtained in the process of fitting the model to the data using an exponential cubic spline (29-33) with 10 nodes evenly distributed over the study period. Note that the time varying basic reproductive number is given by *R*_*0*_ (*t*) *=*β (*t*) /γ in the absence of vaccination.

### Modelling vaccination

#### Immunity waning

In the classic SEIR model, the duration time of individuals remaining in each compartment is exponentially distributed. However, the duration for which vaccinated individuals spend in the *V* compartment is unlikely to be exponentially distributed. Therefore vaccinated individuals in our model were passed through a chain of five serial compartments so that the duration in the *V* classes follows the very flexible gamma distribution (34) whose parameters (and thus shape) can be fitted.

In more detail, vaccinated susceptible individuals were divided into three groups:

a. a proportion (η_*1*_) gain long-term protection and move directly to the recovered R class;
b. a proportion (η_*2*_)gain medium-term temporary protection by passing through a series of five *V*_*(k)*_ classes, during which immunity wanes slowly (set by *1/*κ year), and then finally move on to the *S*_*V*_ class. Individuals in *S*_*V*_ are susceptible to breakthrough infections and move to the *I* class through an exposure stage (*E*_*B*_);
c. and the remaining proportion (η_*3*_)gain little protection from vaccination and move to *SV* directly.

In general, the proportions η_*1*_*+*η_*2*_*+*η_*3*_ *= 1*. These parameters are hard to estimate separately only from observational data, and so their values have to be approximated to some extent to match observed characteristics of the vaccine. We set η_*1*_ *= 0*.*1* to emulate that a small fraction of individuals do not lose immunity (at least in the period modeled in this work), as some level of protection remained over long time periods in waning immunity studies (11). We set η_*3*_ *= 0*.*1* as 1 minus the approximate short term vaccine efficacy (∼90%) estimated from the original vaccine randomized control trials (4) (in the other words, the immediate risk of infection shortly after second dose vaccination is 10%). Hence, η_*2*_ *= 0*.*8*.

For a simple model, if there were only a single vaccination compartment, vaccine-induced immunity would wane exponentially. However, for the full model, by adding a chain of vaccination compartments, the waning is modified to follow a more realistic Gamma distribution with parameters that can remove the rapid exponential decay in waning, which reflects the possibility that the vaccine might be highly effective at least over the first months. For five stages, the waning has a Gamma distribution with shape parameter of 5, and the vaccine efficacy can be expressed as:

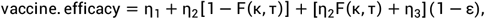

where *F* is the cumulative distribution function of individual protection duration (which is Gamma distributed) for those in the *V* classes (35). The parameter ε reflects the susceptibility of vaccinated individuals to BTI. When ε = 1, vaccinated individuals are as susceptible to infection as unvaccinated. In contrast, when ε = 0, vaccinated individuals have permanent immunity and can never be infected. The parameter τ is the time elapsed since the second dose vaccination. Initially τ *=0*, and the efficacy is η_1_ + η_2_ + η_3_(1 − ε), which over time decreases to a level of η1 + [η_2_ + η_3_](1 − ε) for large τ. Under our parameter values, these two levels are 92% and 28% for example parameters ε = 0.8 (for large τ). In our focal model, with parameters ε = 0.8 and κ^−1^ = 4.28 months where we use five compartments for *V*, the vaccine efficacy also begins at 92% and a calculation shows it will decline to 39% in the six remaining months of the study period. This corresponds to the vaccine efficacy of 39% reported by Israel’s MOH in July 2021 some five months after the second vaccine program was initiated and vigorously pushed out.

When vaccination is modelled with a single stage and thus exponentially distributed (i.e. F = 1 − e^−κτ^), a simpler more intuitive form is obtained

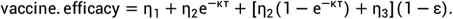

### Technical considerations

Several additional technical modelling considerations have to be dealt with and these are discussed in SI2. For example, the model also assumes that there is a 14-day delay between the date of the second vaccination dose delivered and the onset date of the protective effect. This is implemented by incorporating a time delay whereby the vaccination rate 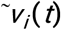 calculated from the data (see SI2) is updated and replaced by 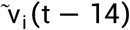. The reduced transmissibility of BTIs, as compared to non-BTIs, is discussed in SI2.

### Model fitting

The plug-and-play likelihood-based inference framework which was implemented in the R package POMP(36, 37) was used. The model was run using the Euler-multinominal integration method, which takes into account demographic noise. A negative-binomial measurement model was used to link model simulated weekly cases and observed weekly cases to take into account measurement noise. These methods have been more thoroughly described in (38, 39).

## Results and Discussion

### Epidemic dynamics captured by the data

In Figure 3, we present the reported weekly confirmed COVID-19 cases in Israel, divided into vaccinated and unvaccinated, for age groups <60 years (panel a) and 60+ years (panel b). Superimposed on the figure is the vaccine coverage for the first, second and third dose, respectively. A more detailed breakdown into age-classes is given in Figure S1. The vaccination program was initiated close to New Year 2021. The number of reported cases increased significantly from December 2020 to January 2021, reaching the largest number of daily infections ever experienced in Israel, when the epidemic rapidly declined from January until June. The country’s mitigation and lockdown policies could have played a large role in the epidemic crash. However, the crash was also simultaneous with the first distribution of the vaccine, which began in Israel in December 2020, initially targeting the elderly population (40). The latter helps to explain why the epidemic curve for the >60 population appeared to respond more quickly, as it decreased over January. Thus, the vaccination program is likely to have been at least partially responsible for the demise of the outbreak.

**Figure 3.**
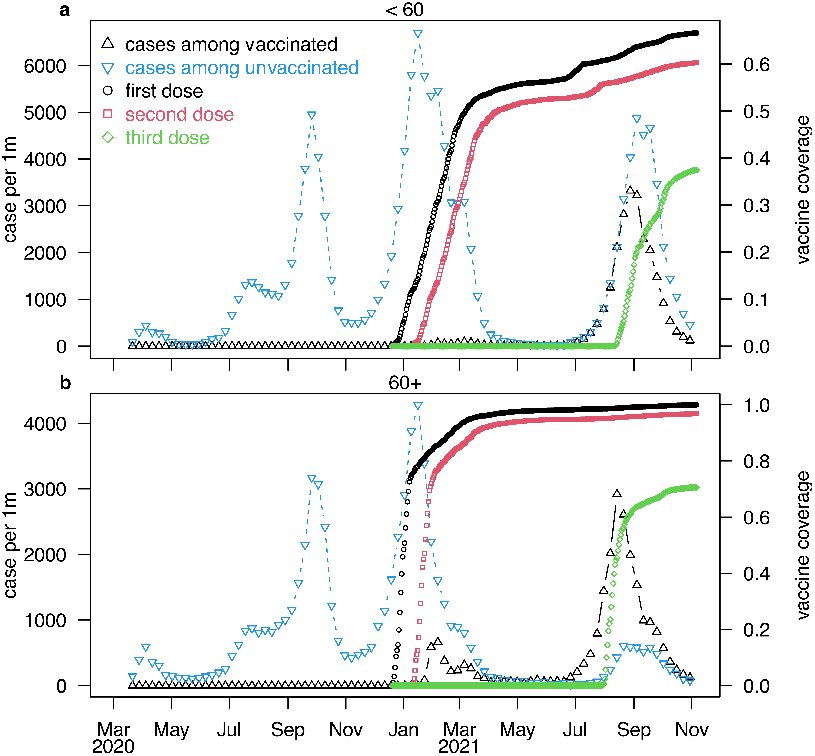
Infection and vaccination dynamics in Israel. Weekly reported COVID-19 cases are presented for the <60 (a) and 60+ (b) age groups (population standardized, per 1 million individuals in an age group), in unvaccinated (blue, down-facing triangles) and fully vaccinated (black, up-facing triangles) individuals. showed The cumulative vaccine coverage for the first, second and third dose are represented by black circles, red squares, and green diamonds. respectively.

By July 2021, more than 60% of the Israeli population were fully vaccinated. Moreover, the country had already experienced three major waves of the pandemic and thus reached a large total attack rate by this point in time. Taken together, these two facts imply that the number of susceptible people remaining in Israel should have become quite limited, explaining why the disease appeared to have become almost eradicated. Despite the appearance of eradication, in late June a surprising fourth wave struck Israel. The data suggest that the two key factors responsible for the fourth wave are *i)* the simultaneous appearance of the highly transmissible Delta variant, which had only just then arrived in Israel; and *ii)* the large numbers of BTIs that began to appear, either due to the waning of immunity or Delta variant vaccine escape.

### Fitting the outbreak dynamics

Here we use the focal model to investigate the above dynamics by fitting the observed epidemic data over the full study period. Specifically, the model is used to fit the four time series seen in Figure 4: infected cases amongst those who were vaccinated (i.e., BTI’s) and infected cases amongst those who were unvaccinated, for two age groups. The vaccination uptake delivered over the first two doses 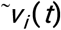 was determined from Figures 3a&b. Before the third dose, the relative susceptibility of vaccinated *S*_*V*_ individuals was set to ε *= 0*.*8*. The third dose was given only to the second dose receivers, but since many of them appear in several compartments simultaneously, it was modelled simply by assuming a reduced susceptibility by a factor ε for <60 age group and *0*.*5*ε for 60+ age group, which appeared after a 26-day delay (to account for the time to its cumulative coverage). Such a reduction reflects the greater difficulty of vaccinated individuals to contribute to BTI’s. From this point on, the susceptibility to BTI ε was determined by fitting the model to the observed data. In order to reduce parameters, we assume that the relative susceptibility was ε for <60 age group, and *0*.*5*ε for 60+ age group, since the third dose coverage was two-fold higher among 60+ age group than among <60 age group over the study period (Figure 3).

**Figure 4.**
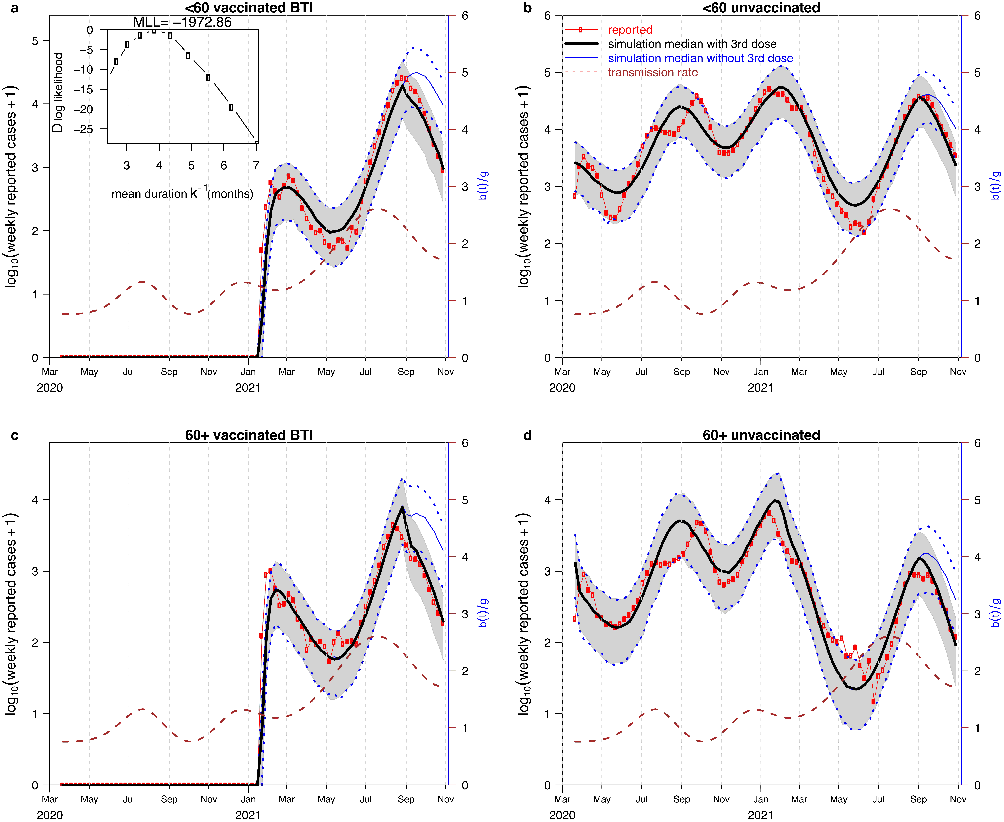
Model fitting results. The mean simulated cases (black bold) versus observed cases (red circles) for the <60 age vaccinated group in (a); for the <60 unvaccinated group in (b); for the 60+ vaccinated group in (c) and for the 60+ unvaccinated group in (d). Note that a logarithmic scale is used. The cases in (a,c) are BTIs. The brown dashed curve shows the transmission rate. The inset in panel (a) shows the profile maximum log-likelihood as a function of κ, and used to determine the best estimate of κ The blue thin curves show the counterfactual scenario where the effect of the third dose is shut off by fixing ε *=0*.*8* constant. The two dotted blue curves show the 95% range of the 1000 simulations when the effect of third dose is shut off and it is possible to see infective numbers in this scenario (Scenario 2).

The model fits are displayed in Figures 4 and 5. The simulated case numbers provide an excellent fit to the reported case numbers in all four groups: <60 vaccinated (a), <60 unvaccinated (b), 60+ vaccinated (c), and 60+ unvaccinated (d), over all four waves of the epidemic. Note that in Figure 4, a logarithmic scale is used to provide a clearer statistical presentation of the fits, while Figure S2 shows the untransformed graph. The reconstructed transmission rate *R*_*0*_ *t =*β(*t*)/γ is plotted as a brown dashed curve, and is the time-varying basic reproductive number (i.e., what it would be if vaccination were absent). The transmission rate *R*_*0*_(*t*) peaked in July 2020 and December 2020, and then decreased. Presumably, the peak in December can be attributed to kindergartens and schools being gradually reopened since the end of October, while the decrease is plausibly due to the restrictions of social distance and nation-wide lockdown implemented by the Israeli government (41-43). In June 1, 2021, Israel revoked almost all the restrictions, apart from mask wearing indoors, and transmission rate kept increasing (44). We note that our estimated *R*_*0*_(*t*) is consistent with the R_t_ empirically estimated by the Israeli MoH until vaccines were deployed in Israel, when the estimates diverge and R_t_ becomes substantially lower. The correspondence between the two estimates is shown in Supplementary Figure S6.

**Figure 5.**
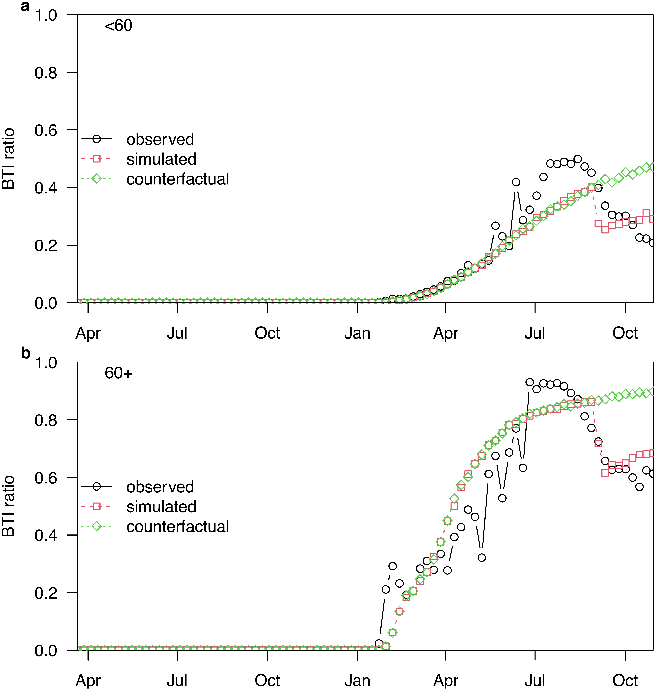
The observed (black circles) versus the simulated BTI ratios under factual (red squares) and counterfactual (green diamonds) scenarios for <60 age group (a) and for 60+ age group (b). These results correspond to Figure 4, showing the BTI ratio instead of case numbers.

Finally, the time varying transmission rate R_0_ t plotted in Figure 4 (dashed line) gives an indication of the transmissibility of the Delta variant. In particular it can be seen that the peak transmission rate after the Delta invasion is 1.96 times larger than before the invasion.

In Figure 5, we show the BTI ratio (BTI cases out of all cases for each age group), based on the case numbers in Figure 4.

Through the model fitting, the temporary duration of protective immunity in the vaccination, or “zero susceptibility” stage, of those for whom the vaccine is effective but wanes (the fraction η_2_), was estimated as κ^−1^ =1/3.096= 3.876 months (Figure 4.a, inset). The vaccine efficacy begins at 92% in January 2021, and the model fit determined it declined to 35% in the six remaining months of the study period. This is comparable to estimates from the literature where immunity is found to wane substantially between two and six months (11) after second dose vaccination, and reports of reaching 39% have been published (15).

In comparison to the susceptibility set at ε =0.8 for the period before the third dose of vaccination, after initiation of the third dose the model fit yielded ε =0.371 for <60 age group and ε =0.186 for 60+ age group. Thus, as a result of the booster, the susceptibility of vaccinated cases is considerably reduced by more than 50%. This echoes the fact that the third dose appears to be very effective in various reports (5, 17)

The reporting ratios of cases ρ_*i*_ were found to be approximately 48.3% and 50.5% for two age groups, respectively, which are epidemiologically reasonable. Namely, for any reported case, there was an additional unreported case.

Of particular interest is that the Delta variant invaded Israel some 4-5 months after the second dose of the vaccine was distributed to the population on a large scale. With the waning of immunity, the Delta variant outbreak increased the BTI ratio which peaked at 85% for the 60+ age group, in the end of August 2021, and then dropped sharply. This can be contrasted with the BTI levels observed if assuming that the susceptibility would have remained at the same level with ε =0.8 (see Figure 5).

We argue that this drop may be related to the implementation of the third dose of the vaccine. As mentioned above, we assumed that before August 25, 2021, the reduced relative susceptibility of the vaccinated was ε *=0*.*8* and then, with the passage of time, it dropped to ∼0.3 (estimated from the data), which led to the drop in the BTI ratio. Together, these two effects, invasion of the Delta variant and implementation of the third dose, account for the observed BTI pattern. However, the exact contribution of immunity waning and the increased transmissibility, or immune escape, of the Delta variant is difficult to disentangle given the current data.

### Breakthrough infections

We use the breakthrough infection (BTI) ratio as a key index in this work, and expand on this further here. The BTI ratio is the ratio of the number of BTIs (fully vaccinated individuals that become infected) among the total infected, at a given point in time for an age group. It is defined for the *i*’th age-group as:

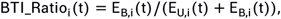

where *i=0,1* represents populations <60 and >60 years of age, respectively; *E*_*B,i*_ is the number of exposed cases due to BTI, i.e. among the vaccinated; and *E*_*U,i*_ is the number of exposed cases among the unvaccinated. BTIs generally lead to milder symptoms than infections amongst the unvaccinated in the same age-group (45, 46). Thus, the BTI ratio among all severe cases, denoted as *BTI_Ratio*_*i*__severe, should be smaller than the BTI ratio amongst all infected individuals. That is, *BTI_Ratio*_*i*__severe < *BTI_Ratio*_*i*_. Figure 6 shows the BTI dynamics during August 2021. In Figure 6.a, we can see that *BTI_Ratio*_*i*__severe in the 60+ age group is greater than 60% for most of August 2021. As mentioned above, since BTI cases should have relatively mild symptoms, the BTI ratio amongst all infections (both severe cases and mild cases) should be no less than 60% among the 60+ age-group.

**Figure 6.**
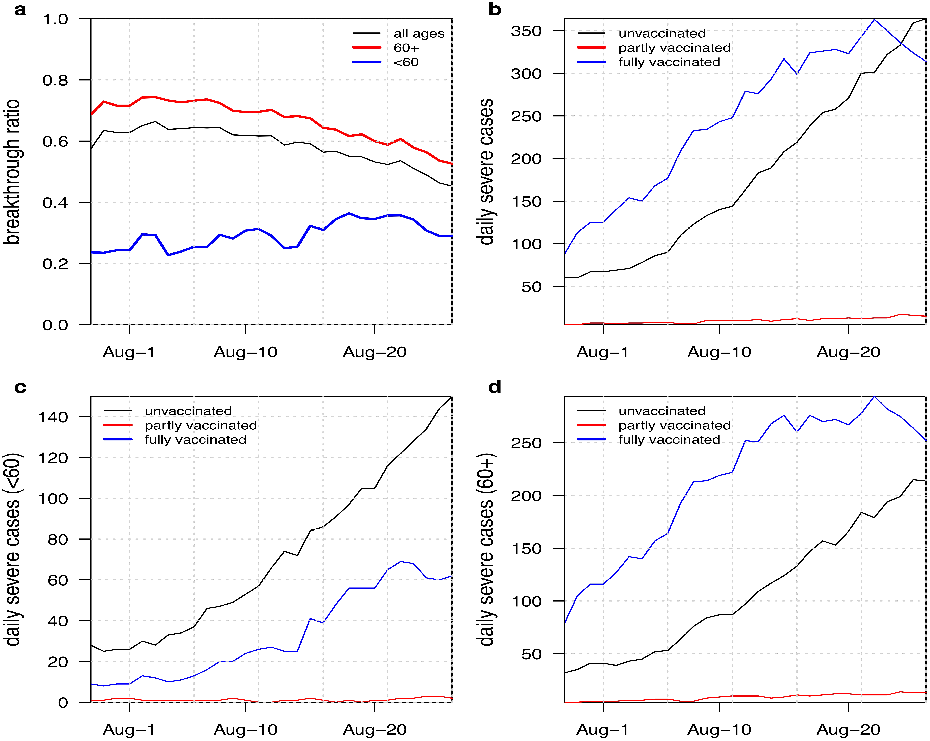
(a) Breakthrough infection ratio among severe cases (*BTI_Ratio*_*i*__severe) over time for all ages, age 60+ and age <60. (b) Daily reported severe cases for three types (unvaccinated, partly vaccinated, fully vaccinated) for all ages over time. (c) three types of daily severe cases for <60 years old. (d) three types of daily severe cases for 60+ year olds.

We observe that the BTI among all cases in the 60+ group reached 85%. Furthermore, there were about 60% BTIs among severe cases in August 2021 (Figure 6). In other words, if *n* individuals were infected, *0*.*85n* would have been vaccinated. If a proportion *q* of the infected developed a severe disease, *0*.*6q* of them would have been vaccinated. Hence the relative risk of severe outcome among BTI compared with unvaccinated is 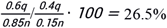. Therefore, given a breakthrough infection, vaccine effectiveness is 73.5% against severe outcome.

### Counterfactual model with booster “switched off”

We generated simulations from a counterfactual model, in which the third booster was removed or “switched to zero”. This was achieved by running the focal model with the same parameters (including the same *R*_*0*_ (*t*)) apart from the susceptibility ε, which was kept fixed to ε = 0.8 for the entire time period, as if there were no third vaccination. The model thus simulated the number of infectives expected in the absence of the booster. The simulated time series in this scenario (Figure 4, blue curves) passes through an epidemic peak larger in magnitude than the observed time series (Figure 4, red curves), especially for the 60+ population.

The cumulative number of infectives can be compared with that of the focal model to find the number of cases averted by the third dose vaccination. Under this scenario, we estimated that the third dose vaccination prevented approximately 469,680 (95%CI 195,729, 766,710) reported cases. A clearer visual representation of this scenario is presented in Figure S8 which plots the time series of Figure 4 without the logarithmic scaling.

Considering a reporting ratio of 50%, the actual number of infections prevented could have been approximately 939,360. We emphasize that this counterfactual scenario should be interpreted with caution, as it assumes that all other factors would have remained constant. It is likely, however, that a sharp increase in infections would have led to more governmental restrictions, in turn reducing infection rates.

### Alternative Scenarios

We examined and analysed several alternatives to our focal model as sensitivity checks. First, we created a model that assumes the booster was never given, by fixing ε *=0*.*8* throughout the entire study time-period (see supplementary SI3 Scenario2). The model (lacking the effect of the booster) was then fitted to the full observed data set of the Israeli population that had experienced a booster. A failure of the model to fit the data well indicates that the actual booster has modified the population’s epidemic dynamics. Indeed, in supplementary Section SI3 and Figure S3 we show that without the third dose the model has a worse fit, decreasing by 24.18 log-likelihood units or a Δ*AICc=46*.*12* (the second order Akaike Information Criterion), which indicates a significant difference (47).

We further fitted the focal model, but with only a single stage in the *V* class, and thus an exponentially distributed duration of vaccination. In supplementary Section SI3 (Scenario 3) and Figure S4, we show that under this assumption, the model fit is decreased by 15.87 log-likelihood units and a Δ*AICc=31*.*737* which is a significant difference. Thus, the Gamma distributed duration of vaccine-induced immunity protection employed in the focal model is preferable to an exponentially distributed duration of protection.

Thus the third dose effects and the Gamma distributed duration of vaccine-induced immunity protection are necessary to properly fit the model to the data.

### Conclusions

In summary, we propose a model framework analyzing COVID-19 dynamics in Israel and shedding light on important issues of BTI and waning immunity. Through mathematical modeling, we showed that the fourth wave of COVID-19 in Israel was a result of low vaccination coverage among <60 years individuals, the large proportion of BTI among vaccinated age 60+ years individuals (reaching levels of 85%), and a compromised vaccine efficacy due to a combined effects of immunity decay and the invasion of the Delta variant. Through a model reconstruction of the reproductive number R0 (*t)*, it was found that the peak transmission rate of the Delta variant was 1.96 times larger than the previous Alpha variant. In addition, the model found that either the vaccine induced immunity dropped significantly or the Delta variant possesses immunity escaping abilities. Thus vaccine efficacy began at 92% in January 2021 but the model estimated it must have dropped to 39% 5-6 months later. We also presented a BTI analysis, showing that the BTI ratio indeed peaked during the Delta strain invasion, and before the third booster.

Furthermore, we simulated a counterfactual scenario where the third vaccine dose, or booster, was precluded. Under such a scenario, hundreds of thousands of new infections would have occurred, had there been no further governmental interventions. Hence, the significance of the booster in mitigating the Israeli COVID-19 epidemic cannot be overstated.

Our analyses were applied to Israel – which has a highly vaccinated population, and an effective health care system with an efficient and publically available data collection system. It therefore served as a perfect case study, especially given its early drive to hastily redistribute the booster dose. Thus the lessons learned from the dynamics in Israel may serve to increase preparedness in other locations observing invasions of variants of concern.

In the future, our proposed framework can also be easily applied to new data on emerging variants, to compare and examine different scenarios as this epidemic or other outbreaks unfold.

## Data Availability

All data produced in the present study are available upon reasonable request to the authors.
All data produced in the present work are contained in the manuscript.
All data produced are available online at the websites of Israel Ministry of Health (MOH) and the World Health Organization (WHO).

https://data.gov.il/dataset/covid-19/resource/89f61e3a-4866-4bbf-bcc1-9734e5fee58e?inner_span=True

https://data.gov.il/dataset/covid-19/resource/e4bf0ab8-ec88-4f9b-8669-f2cc78273edd?inner_span=True

https://data.gov.il/dataset/covid-19/resource/57410611-936c-49a6-ac3c-838171055b1f?inner_span=True

https://datadashboard.health.gov.il/COVID-19/general?tileName=SeriousVaccinationStatusDaily

https://ourworldindata.org/grapher/covid-variants-area?country=~ISR

## Declarations

### Availability of data and materials

All data used in this work were publicly available. The data source is obtained in: https://datadashboard.health.gov.il/COVID-19/general

## Acknowledgments

We thank Stan Yip, Shi Zhao and Yinou Wang for insightful discussion.

## Contributions

All authors conceived the study, carried out the analysis, wrote the draft, revised the manuscript critically, and approved it for publishing.

## Ethics declarations

### Ethics approval and consent to participate

Not applicable.

### Consent for publication

Not applicable.

---

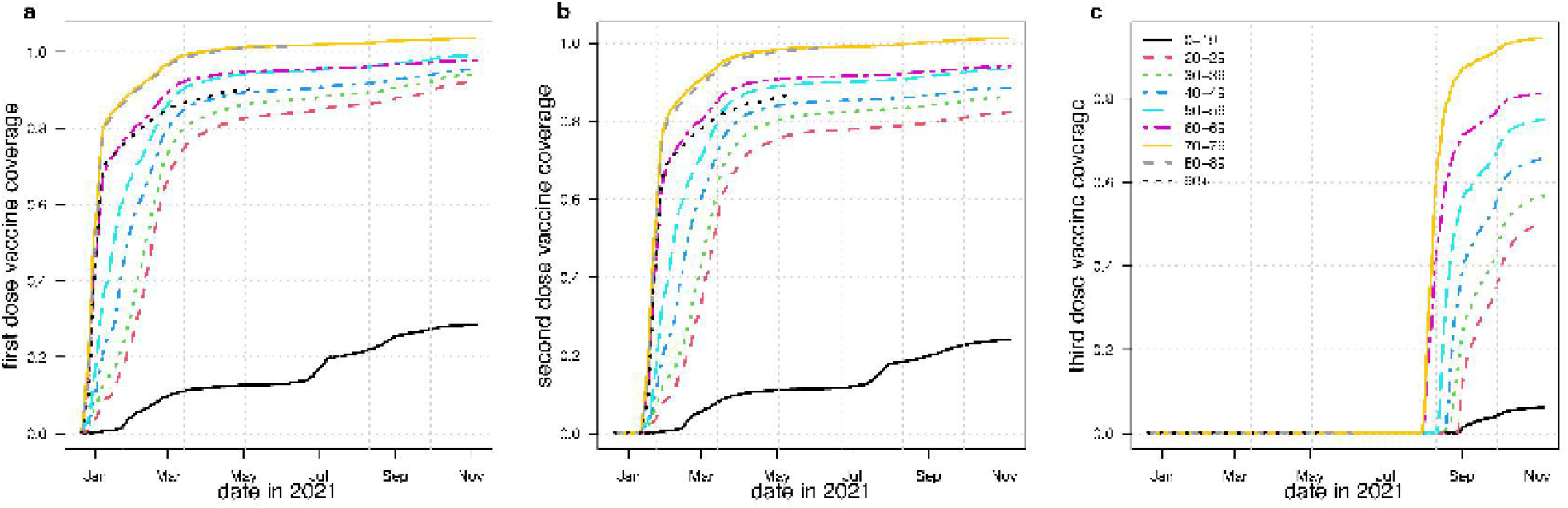

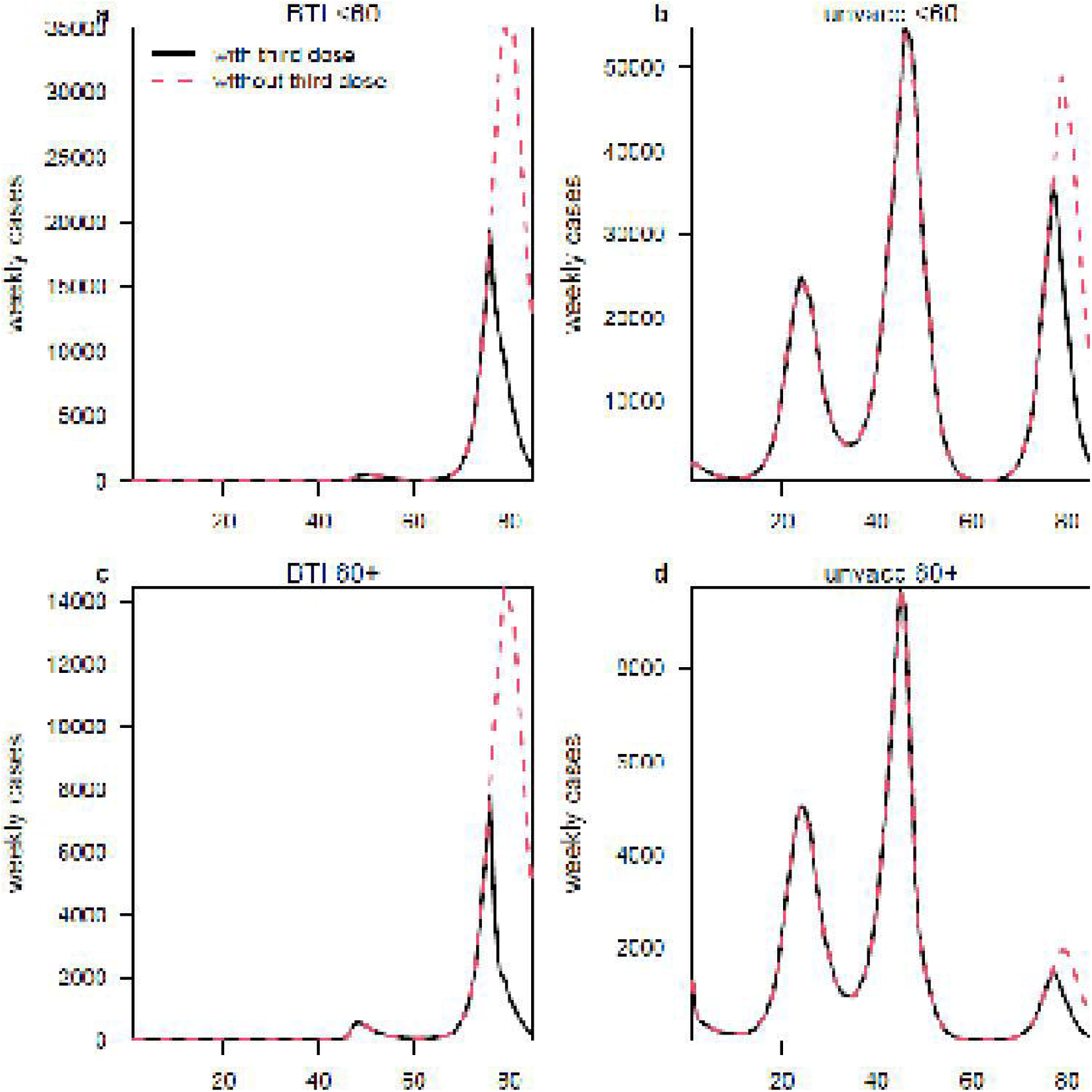

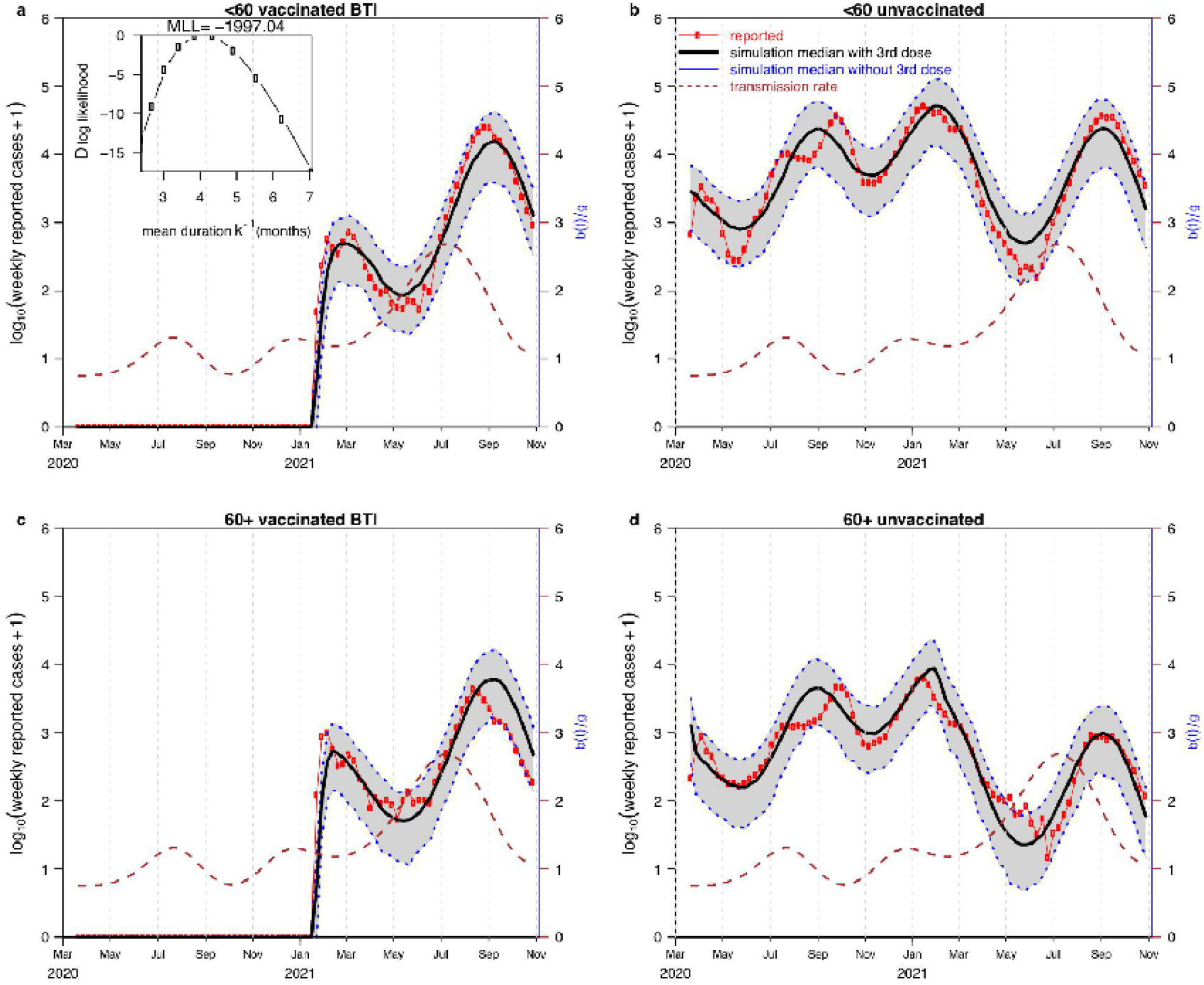

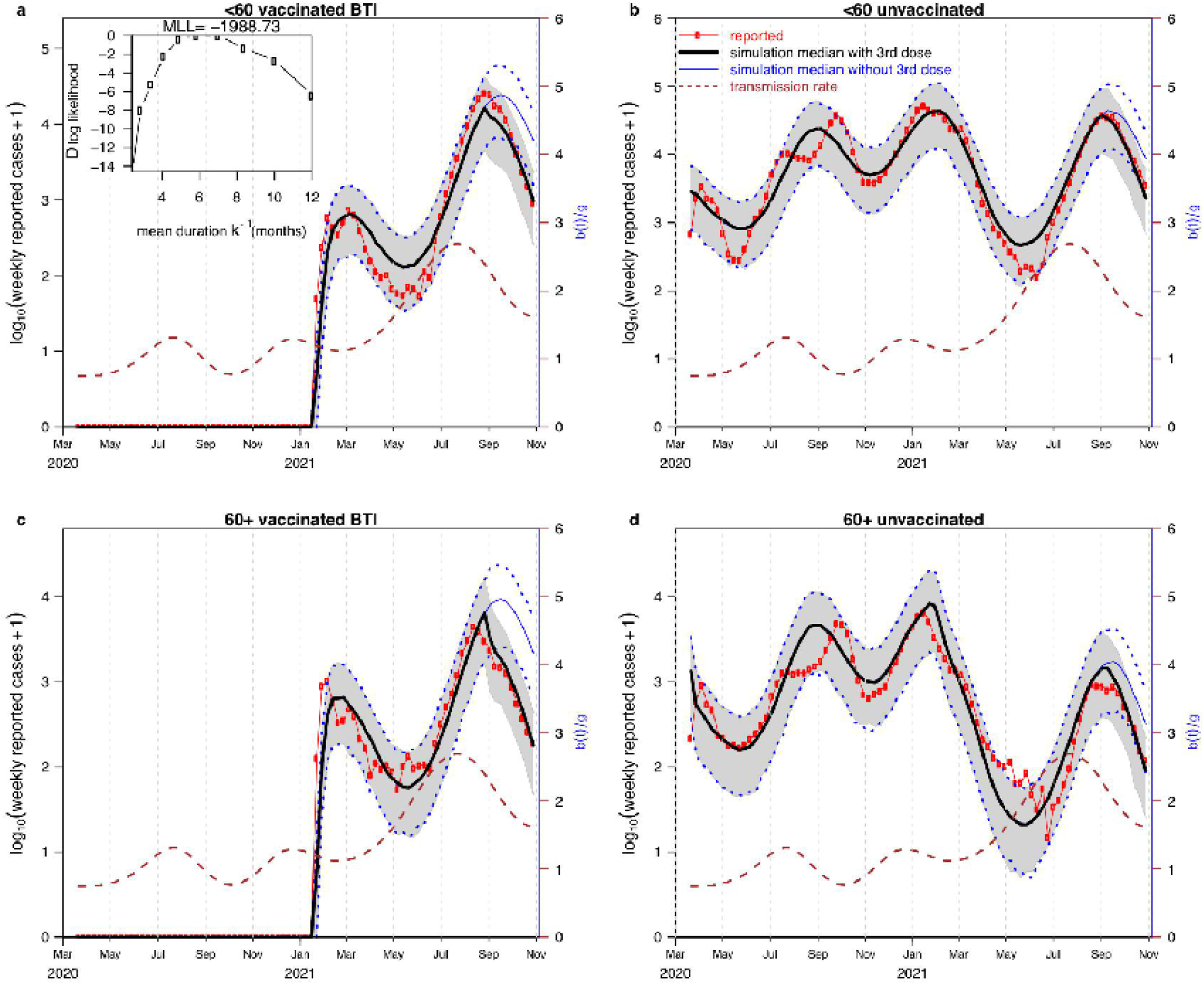

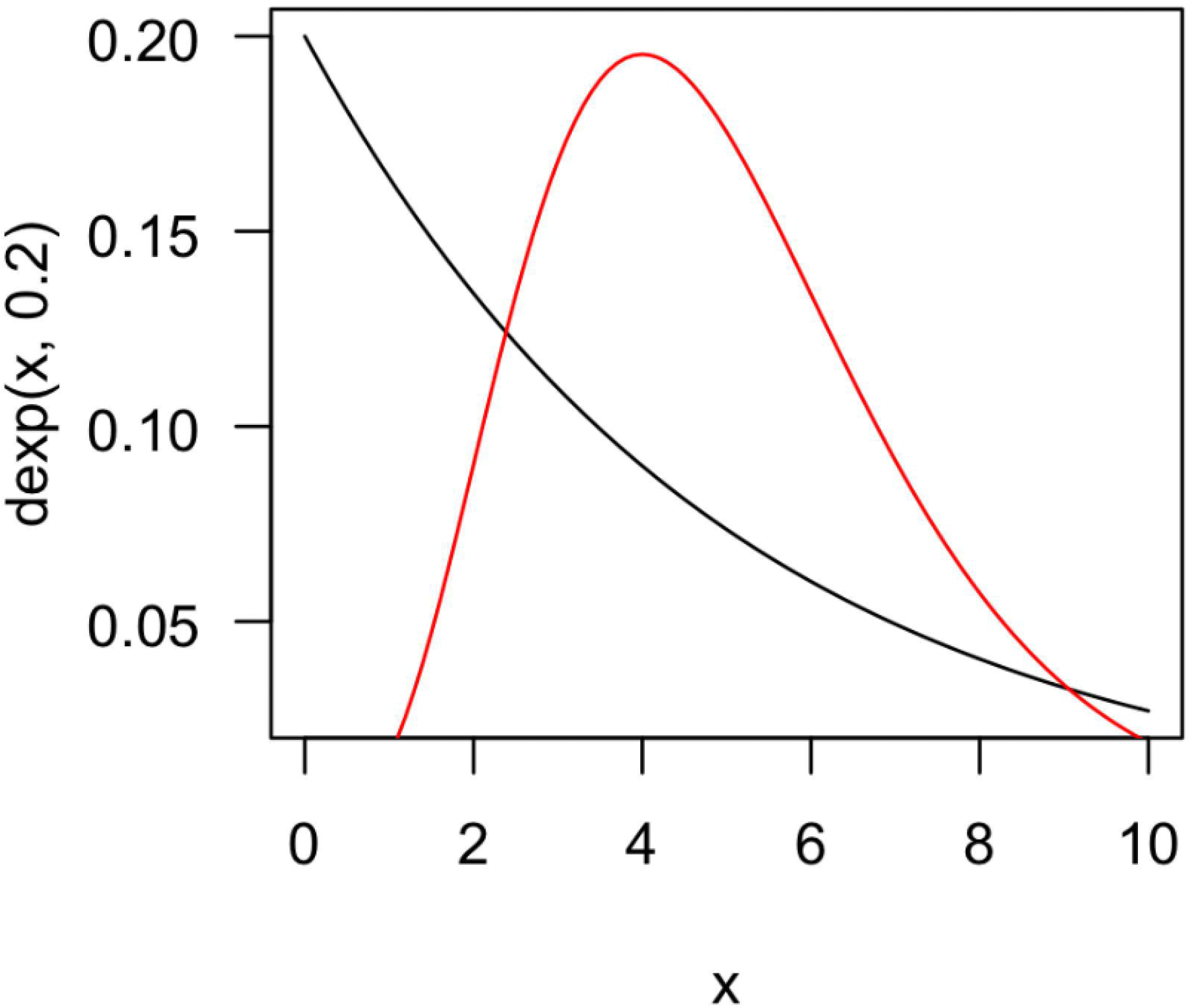

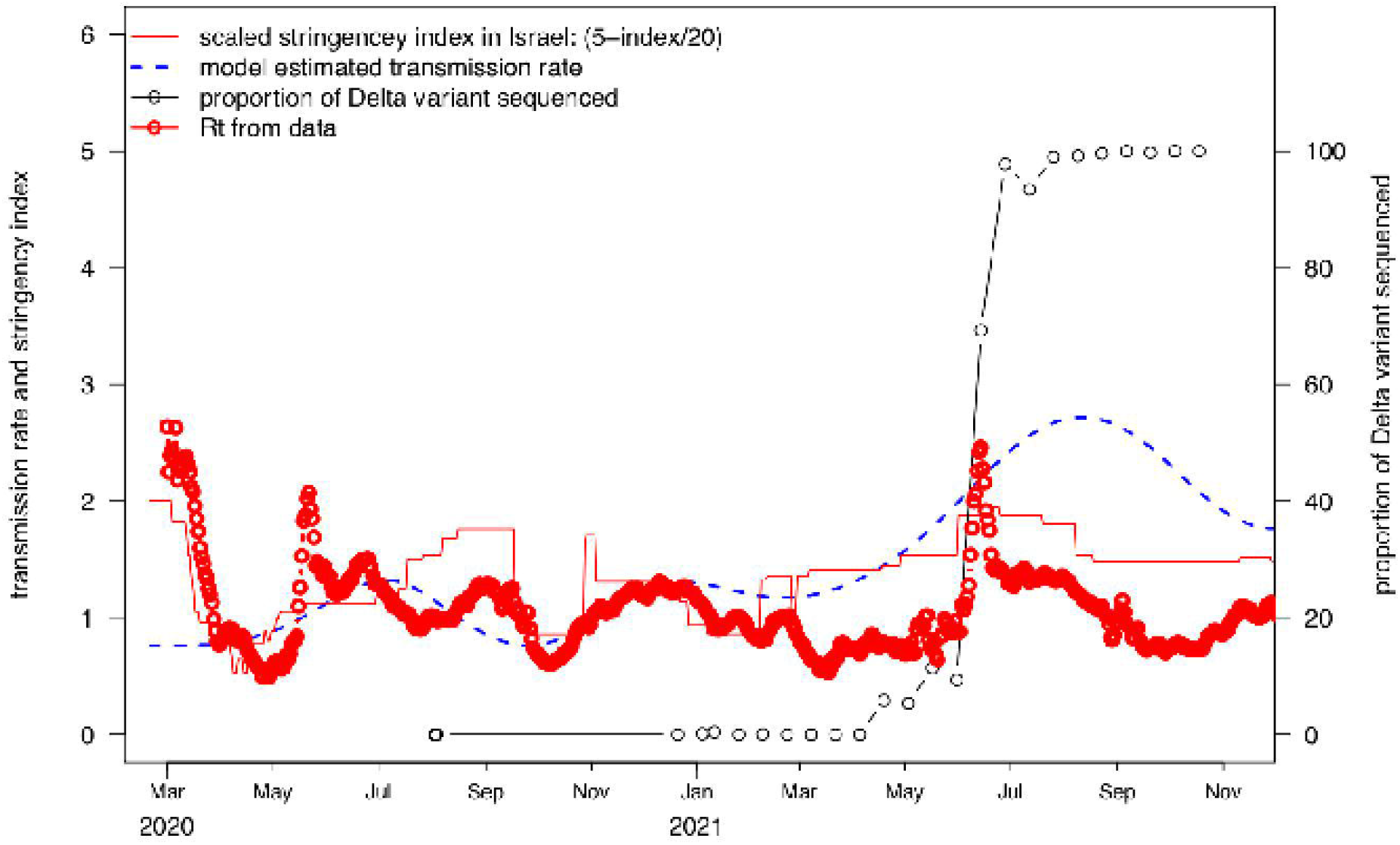

## References

1. Mlcochova P, Kemp SA, Dhar MS, Papa G, Meng B, Ferreira IA, et al. SARS-CoV-2 B. 1.617. 2 Delta variant replication and immune evasion. Nature. 2021:1–6.

2. Israel urges vaccination for all teens, citing Delta variant.Reuters website.

3. Mallapaty S. Will COVID become a disease of the young? Nature. 2021;595(7867):343–4.

4. Polack FP, Thomas SJ, Kitchin N, Absalon J, Gurtman A, Lockhart S, et al. Safety and efficacy of the BNT162b2 mRNA Covid-19 vaccine. New England Journal of Medicine. 2020.

5. Vogel G. Israel’s COVID-19 boosters are preventing infections, new studies suggest. Science. 2021.

6. Definition of Variants of SARS-CoV-2. Centers for Disease Control and Prevention website [Available from: https://chinese.cdc.gov/coronavirus/2019-ncov/variants/variant-info.html.

7. Israel to reimpose coronavirus ‘Green Pass’ as Delta variant hits: Reuters; 2021 [Available from: https://www.reuters.com/world/middle-east/israel-reimpose-coronavirus-green-pass-delta-variant-hits-2021-07-22/.

8. Israel extends lockdown, sees delay in COVID-19 turnaround: Egypt Independent; 2021 [Available from: https://egyptindependent.com/israel-extends-lockdown-sees-delay-in-covid-19-turnaround/.

9. Israel: Authorities reimpose domestic COVID-19 restrictions until Aug. 8 due to increase in cases: GARDAWORLD; 2021 [Available from: https://www.garda.com/crisis24/news-alerts/507946/israel-authorities-reimpose-domestic-covid-19-restrictions-until-aug-8-due-to-increase-in-cases-update-111.

10. Haas EJ, Angulo FJ, McLaughlin JM, Anis E, Singer SR, Khan F, et al. Impact and effectiveness of mRNA BNT162b2 vaccine against SARS-CoV-2 infections and COVID-19 cases, hospitalisations, and deaths following a nationwide vaccination campaign in Israel: an observational study using national surveillance data. The Lancet. 2021;397(10287):1819–29.

11. Goldberg Y, Mandel M, Bar-On YM, Bodenheimer O, Freedman L, Haas EJ, et al. Waning immunity after the BNT162b2 vaccine in Israel. New England Journal of Medicine. 2021.

12. Kupferschmidt K, Wadman M. Delta variant triggers new phase in the pandemic. American Association for the Advancement of Science; 2021.

13. Fisman T. Age-Specific Changes in Virulence Associated with SARS-CoV-2 Variants of Concern. medRxiv. 2021.

14. Sokol S. Israel’s 17% Unvaccinated Now Account for 65% of All Serious COVID-19 Cases 2021 [Available from: https://www.haaretz.com/israel-news/israel-unvaccinated-booster-65-serious-covid-19-cases-death-delta-1.10208784.

15. Jr. BL. Israel says Pfizer Covid vaccine is just 39% effective as delta spreads, but still prevents severe illness. CNBC. 2021.

16. Decline in vaccine effectiveness against infection and symptomatic illness. Israel Ministry of Health website 2021 [Available from: https://www.gov.il/en/Departments/news/05072021-03..

17. Bar-On YM, Goldberg Y, Mandel M, Bodenheimer O, Freedman L, Kalkstein N, et al. Protection of BNT162b2 vaccine booster against Covid-19 in Israel. New england journal of medicine. 2021;385(15):1393–400.

18. Weekly reported cases stratified by age [Available from: https://data.gov.il/dataset/covid-19/resource/89f61e3a-4866-4bbf-bcc1-9734e5fee58e?inner_span=True.

19. Daily hospitalization condition including mean age, gender, and severity of patients [Available from: https://data.gov.il/dataset/covid-19/resource/e4bf0ab8-ec88-4f9b-8669-f2cc78273edd?inner_span=True.

20. Daily vaccination on three doses stratified by age [Available from: https://data.gov.il/dataset/covid-19/resource/57410611-936c-49a6-ac3c-838171055b1f?inner_span=True.

21. Daily diagnosed severe cases stratified by age [Available from: https://datadashboard.health.gov.il/COVID-19/general?tileName=SeriousVaccinationStatusDaily.

22. Organization WH. Daily confirmed cases and deaths for all ages 2021.

23. Variant of Concern sequenced proportion [Available from: https://ourworldindata.org/grapher/covid-variants-area?country=~ISR.

24. Serious vaccination status daily. Israel Minitry of Health website. [Available from: https://datadashboard.health.gov.il/COVID-19/general?tileName=SeriousVaccinationStatusDaily.

25. Brauer F, Driessche Pd, Wu J. Lecture notes in mathematical epidemiology. Berlin, Germany Springer. 2008;75(1):3–22.

26. Tang X, Musa SS, Zhao S, Mei S, He D. Using Proper Mean Generation Intervals in Modeling of COVID-19. Frontiers in public health. 2021;9.

27. He D, Artzy-Randrup Y, Musa SS, Gräf T, Naveca F, Stone L. The unexpected dynamics of COVID-19 in Manaus, Brazil: Was herd immunity achieved? medRxiv. 2021:21251809.

28. Stone L, He D, Lehnstaedt S, Artzy-Randrup Y. Extraordinary curtailment of massive typhus epidemic in the Warsaw Ghetto. Science Advances. 2020;6(30):eabc0927.

29. Bartels RH, Beatty JC, Barsky BA. An introduction to splines for use in computer graphics and geometric modeling: Morgan Kaufmann; 1995.

30. Vetterling WT, Press WH, Teukolsky SA, Flannery BP. Numerical recipes: example book C (The Art of Scientific Computing): Press Syndicate of the University of Cambridge; 1992.

31. Burden RL, Faires JD. Numerical analysis 8th ed. Thomson Brooks/Cole. 2005.

32. Song H, Fan G, Zhao S, Li H, Huang Q, He D. Forecast of the COVID-19 trend in India: a simple modelling approach. 2021.

33. Song H, Fan G, Liu Y, Wang X, He D. The second wave of COVID-19 in South and Southeast Asia and vaccination effects. 2021.

34. Krylova O, Earn DJ. Effects of the infectious period distribution on predicted transitions in childhood disease dynamics. Journal of The Royal Society Interface. 2013;10(84):20130098.

35. Gamma distribution [Available from: https://en.wikipedia.org/wiki/Gamma_distribution.

36. Ionides EL, Bretó C, King AA. Inference for nonlinear dynamical systems. Proceedings of the National Academy of Sciences. 2006;103(49):18438–43.

37. He D, Ionides EL, King AA. Plug-and-play inference for disease dynamics: measles in large and small populations as a case study. J Royal Society Interface. 2010;7(43):271–83.

38. He D, Zhao S, Lin Q, Musa SS, Stone L. New estimates of the Zika virus epidemic attack rate in Northeastern Brazil from 2015 to 2016: A modelling analysis based on Guillain-Barré Syndrome (GBS) surveillance data. PLoS Negl Trop Dis. 2020;14(4):e0007502.

39. Zhao S, Stone L, Gao D, He D. Modelling the large-scale yellow fever outbreak in Luanda, Angola, and the impact of vaccination. PLoS Negl Trop Dis. 2018;12(1):e0006158.

40. The First Ones to Have the COVID-19 Vaccine within the “Ten Katef” Vaccine Campaign: Ministry of Health; 2020 [Available from: https://www.gov.il/en/departments/news/16122020-01.

41. Third coronavirus lockdown rules - everything you need to know: The Jerusalem Post; 2020 [Available from: https://www.jpost.com/israel-news/coronavirus-third-lockdown-rules-everything-you-need-to-know-653072.

42. Israeli gov’t imposes new restrictions: Weekend lockdowns, closure of gyms, studios: i24 News; 2020 [Available from: https://www.i24news.tv/en/news/coronavirus/1594955861-israel-new-restrictions-impose-weekend-lockdown-close-gyms-studios.

43. Government orders closure of event halls, culture venues, gyms and nightclubs: The Times of Israel; 2020 [Available from: https://www.timesofisrael.com/government-okays-closure-of-event-halls-culture-venues-gyms-and-nightclubs/.

44. Back to normal: Israel lifts nearly all COVID restraints as virus fades away: The Times of Israel; 2021 [Available from: https://www.timesofisrael.com/back-to-normal-israel-lifts-nearly-all-covid-restraints-as-virus-fades-away/.

45. Bergwerk M, Gonen T, Lustig Y, Amit S, Lipsitch M, Cohen C, et al. COVID-19 breakthrough infections in vaccinated health care workers. New England Journal of Medicine. 2021.

46. Risk for COVID-19 Infection, Hospitalization, and Death By Age Group: Center for Disease Control and Prevention, United States; [Available from: https://www.cdc.gov/coronavirus/2019-ncov/covid-data/investigations-discovery/hospitalization-death-by-age.html.

47. Anderson D, Burnham K. Model selection and multi-model inference. Second NY: Springer-Verlag. 2004;63(2020):10.

