## Supplementary Information for "Modelling COVID-19 Vaccine Breakthrough Infections in Highly Vaccinated Israel – the effects of waning immunity and third vaccination dose"

**Section 1 Vaccine coverage for three doses among age groups**

Figure S1 shows the vaccine coverage in Israel from December 20, 2020 to November 6, 2021. The vaccine priority was for the elderly first in December 2020, while at the last stages the government approved all adolescents 11-18 to vaccinate beginning in June 2021 (1, 2), and the children from 5-11 years of age since November 14 2021. The ages of those receiving the booster third dose of vaccination (Figure S1 panel c) were mainly adults between 40 and 69 years old. Compared with this age group, older people and younger people were poorly vaccinated.

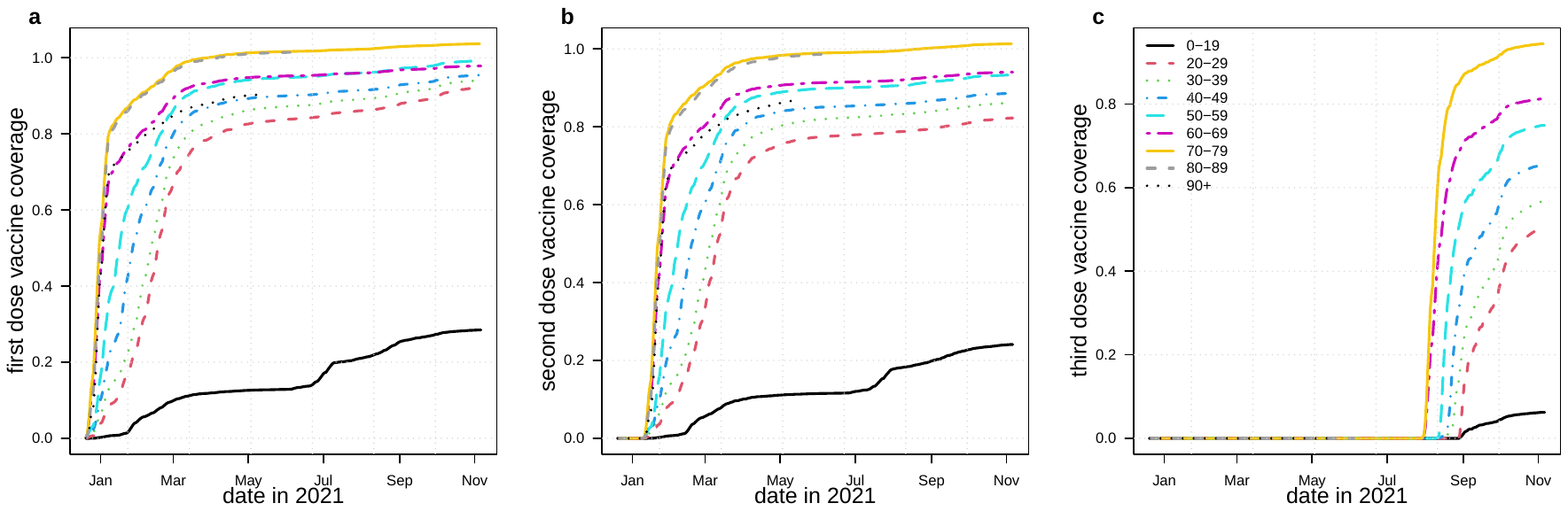

**Figure S1.** Vaccination coverage stratified by age group in three scenarios. (a) LH panel: First dose (partly vaccinated) (b) Middle panel: Second dose (fully vaccinated) (c) RH panel: Third dose. In the coverage in the age group <20 year is much lower than other age groups.

In our model, we model the effects of the third dose by reducing the relative susceptibility of individual in$S_{V}$ on August 25, 2021. The third dose vaccination started on July 30, 2021. On August 25, 2021, the age breakdown vaccination coverage of the third dose is

| Age group | 0-19 | 20-29 | 30-39 | 40-49 | 50-59 | 60-69 | 70-79 | 80-89 | 90+ |
| --- | --- | --- | --- | --- | --- | --- | --- | --- | --- |
| Third dose coverage on August 25 2021 | 0 | 0 | 0.0729 | 0.237 | 0.464 | 0.666 | 0.837 | 0 | 0 |

**Section 2 Technical Issues**

***a) Reduced infectivity of Breakthrough Infection cases.*** The BNT162b2 vaccine reduces the infectivity of vaccinated individuals (47). The reduced infectivity ($\omega<1)$ of BTI cases can be modeled as:

$$\beta\left( I_{U}+{\omega I}_{V} \right)=\beta\left( \frac{E}{E+E_{V}}I+\omega\frac{E_{V}}{E+E_{V}}I \right)=\beta I\frac{E+\omega E_{V}}{E+E_{V}} ,$$

where $I_{U}\mathrm{and}I_{V}$represent infections among unvaccinated and vaccinated, respectively. Thus in model equations (1), we replace $\beta$with $\beta\frac{E+\omega E_{V}}{E+E_{V}}$. We assume $\omega=0.8.$

***b) Vaccinating by targeting the unvaccinated:*** The proportion of the whole population that becomes fully vaccinated each day is denoted $v_{i}\left( t \right)$, for age-group $i$. This rate is easily calculated given the cumulative proportion of the population that have been vaccinated daily. In reality, each age group of the population is divided into two sub groups, those vaccinated and those not yet vaccinated, and only the latter group are eligible for vaccination, where susceptible belong to the latter group. To take this into account, the rate at which susceptible were vaccinated, $\tilde{v}_{i}$, for each age group is given by:

$\tilde{v}_{i}\left( t \right)=v_{i}(t)/(1-\int_{0}^{t-1} v_{i}(s)ds)$,

where the denominator is the proportion of the population that is unvaccinated. This conversion is important, one cannot use $v_{i}(t)$ directly, since the unit of $v_{i}\left( t \right)$ is per capita of the whole population per day, rather than per unvaccinated per day, while the vaccination targets the unvaccinated group. The model also assumes that there is a 14-day delay between the date of the second vaccination dose delivered and the onset date of the protective effect. This is implemented by incorporating a time delay whereby the vaccination rate $\tilde{v}_{i}\left( t \right)$ calculated from the data is updated and replaced by $\tilde{v}_{i}\left( t-14 \right)$.

**Section 3 Sensitive Analysis and Model Selection**

**Scenario 1**

As mentioned in the main text, the $V$ class was split into five stages $V_{(i)},i=1,2,3,4,5$, and the rate of movement between stages was set at 5$\kappa$, thus the total duration in V is $\kappa^{-1}$. Now the stay of individuals in the $V$ stage is Gamma distributed with a mean $\kappa^{-1}$ (unit of time).

Consider 80%, 10%, and 10% of the vaccinated move to $V$, $R$ and $S_{V}$, namely an immediate failure rate of 10%, and 10% long term protection over the study period, and 80% temporary protection. The results for this Scenario 1 are reported in the main text. Figure S2 shows results in Figure 4 plotted without logarithmic transformation.

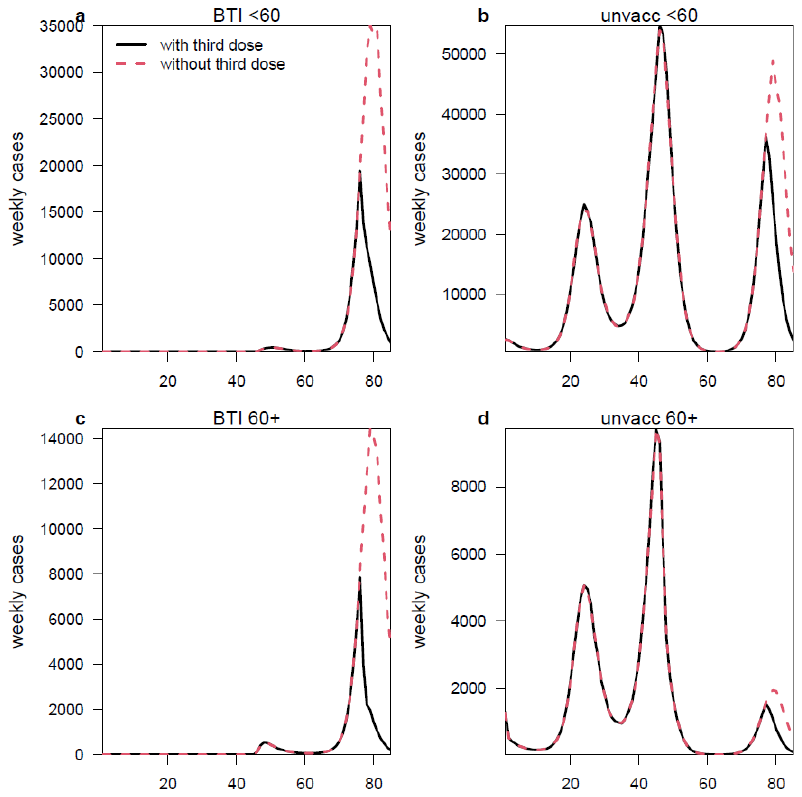

**Figure S2.** Replicate of Figure 4 of main text on normal scale

**Scenario 2**

We performed several alterations to our focal model as sensitivity checks.

First, we created a model that assumes the booster was never given, by fixing $\varepsilon=0.8$ throughout the entire study time-period . The model (lacking the effect of the booster) was then fitted to the full observed data set of a population that in fact experienced a booster. A failure of the model to fit the data well indicates that the actual booster has modified the population’s epidemic dynamics.

Thus the third dose effects and the gamma distributed duration of vaccine-induced immunity protection are necessary to properly fit the model to the data.

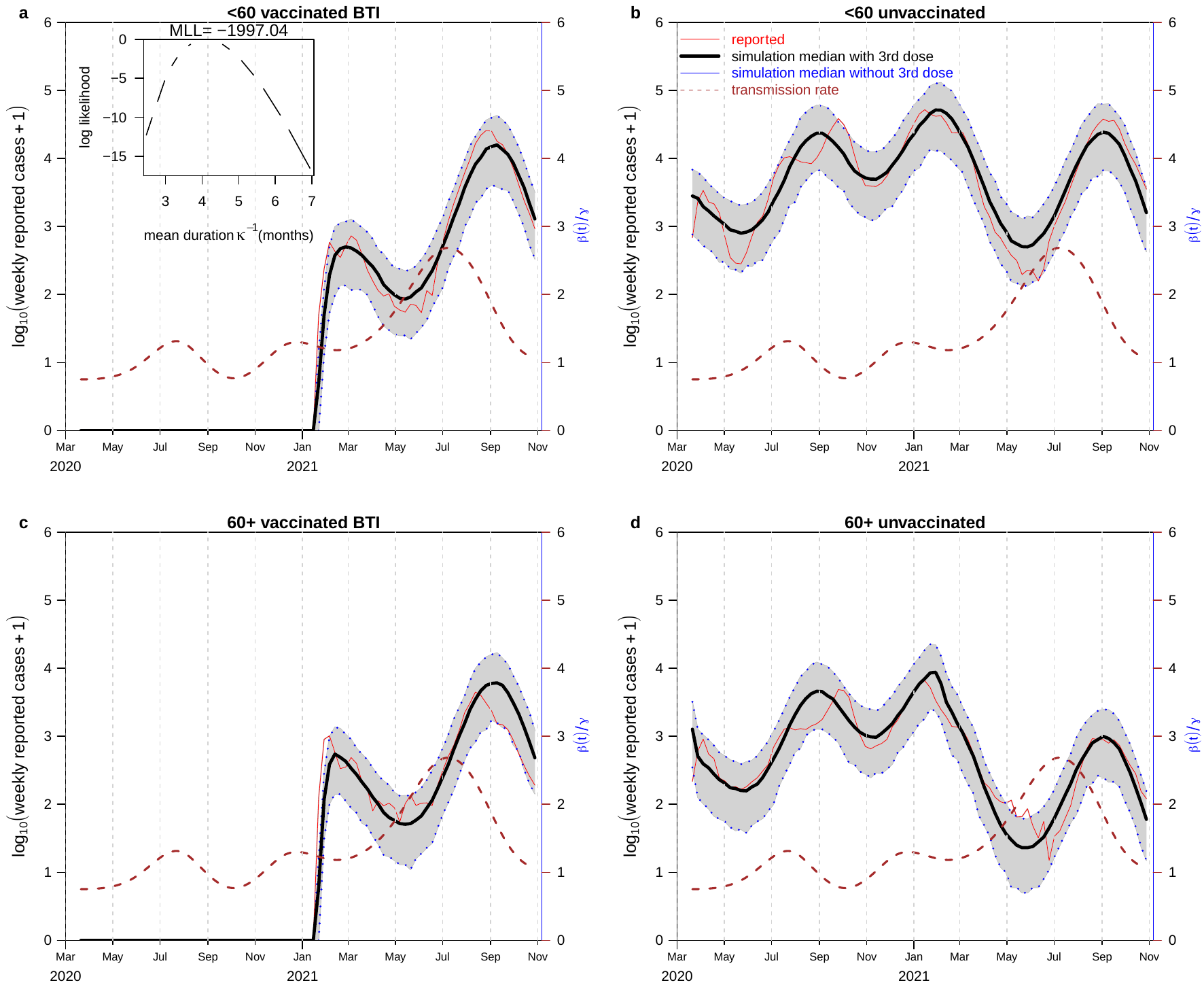

**Figure S3.** A reproduction of Figure 4 with $\varepsilon=0.8$ fixed before and after third dose effects and other parameters are unchanged.

In Scenario 2 Figure S3, we fixed $\varepsilon=0.8$ over the whole study period, the MLL is -1997.04, which is 24 log likelihood units lower than our focal model. This model is significantly worse than our focal model under Akaike Information Criterion.

**Scenario 3**

As another sensitivity check, here we fitted the focal model, but with only a single stage in the V class, and thus an exponentially distributed duration of vaccination.

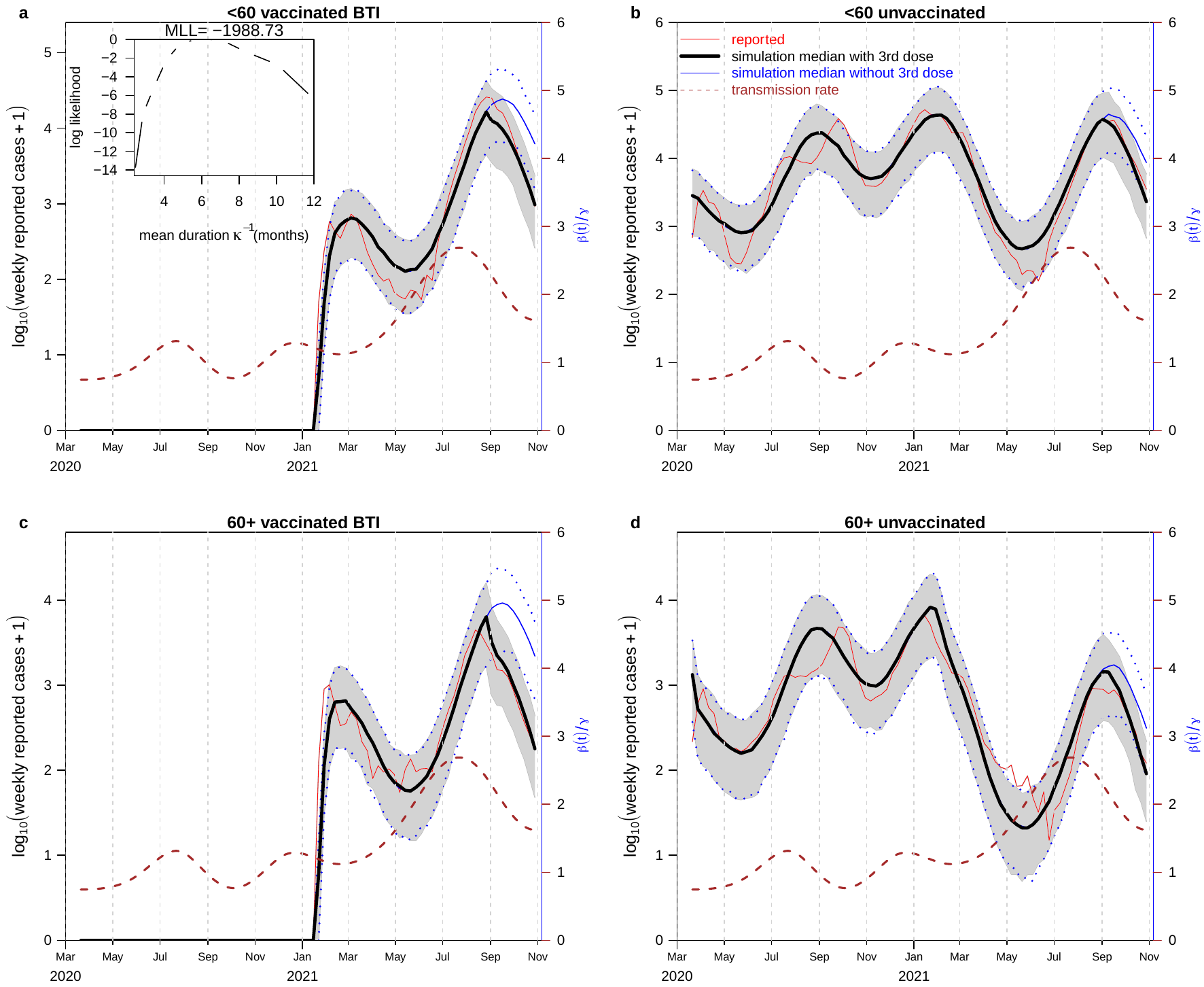

**Figure S4.** Same as Scenario 1, except that a single vaccination class is used giving exponential decay in waning to replace the unimodal Gamma (see Figure S5 below).

In Scenario 3 Figure S4, we find that with an Exponentially distributed model the MLL is 1988.73, which is 15.9 units worsen than our focal model. Thus we demonstrated that the duration of vaccine-induced immunity protection is better modelled as a peaked Gamma distribution (as in the focal model) and having an initial period (the first 2-3 months) in which there is little immunity waning followed by a period of rapid waning.

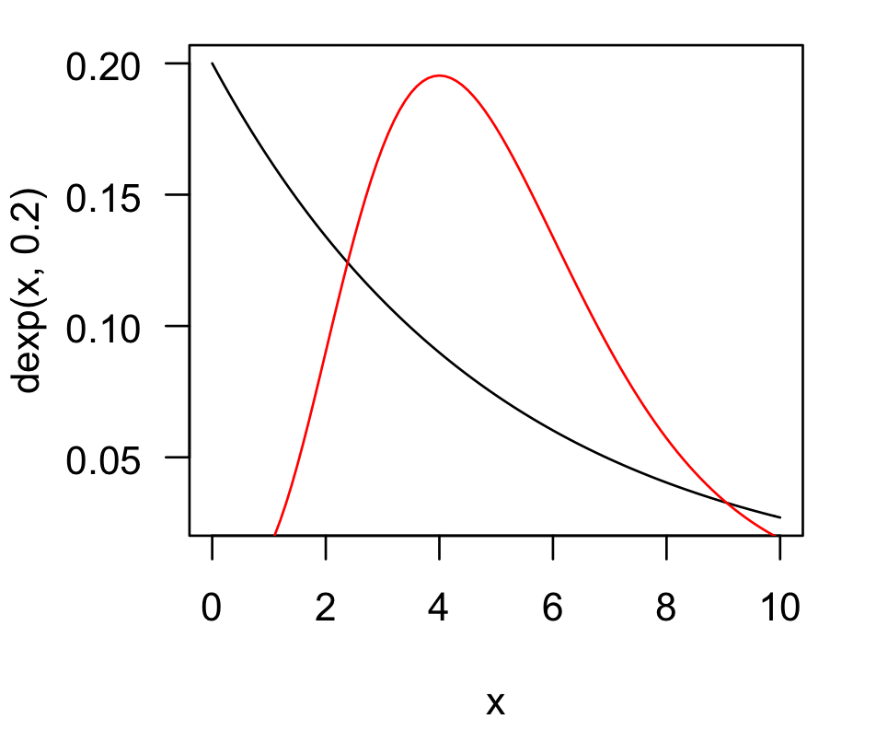

**Figure S5.** A comparison of an Exponential distribution and a Gamma distribution with the same mean.

**Section 4 Stringency Index**

The stringency index composes 9 response indicators to measure the strictness of the policies issued in response to the Covid-19 (4). The response indicators are: workplace closures, school closures, restrictions on public gatherings, cancellation of public events, stay-at-home requirements, closures of public transport, public information campaigns, international travel controls, and restrictions on internal movements. The index value is scaled from 0 to 100, and the higher score reveals the stricter policy response(4).

The calculation of stringency index is described in equation ,

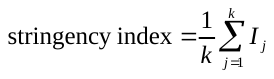
,

where k is the number of indicators and equals to 9 in this equation, and
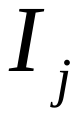
 is the sub-score of each indicator (5).

The sub-score of the indicators are calculated by equation,

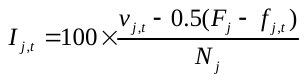
,

where
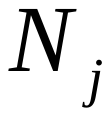
is the maximum value of indicator j,
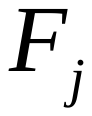
 is the binary flag variable(
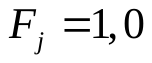
),
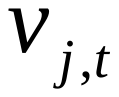
 is the recorded policy value on given day t, and
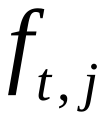
 is the recorded binary flag. Some indicators such as school closures and workplace closures etc. are along with binary flag variable which equals to 1 or 0 (5). The flag variable is related to extra information like geographic scope of certain policy, sectoral scope of revenue support and the funded vaccination by government or individuals(5). Details of the indicator’s values and variables are shown in the codebook of Oxford Covid-19 Government Response Tracker (6). Figure 6 compares the stringency index and our estimated transmission rate, the MOH reproductive number, and the proportion of Delta variant. The estimated transmission rate is equivalent to a time-varying basic reproductive number, which explains the difference between it and the MOH Rt. The proportion of Delta drove the transmission rate to a high value, which is not fully reflected in the changes in the stringency index. Namely the relaxation of stringency and the invasion the Delta strain together increased the transmission rate in Summer 2021.

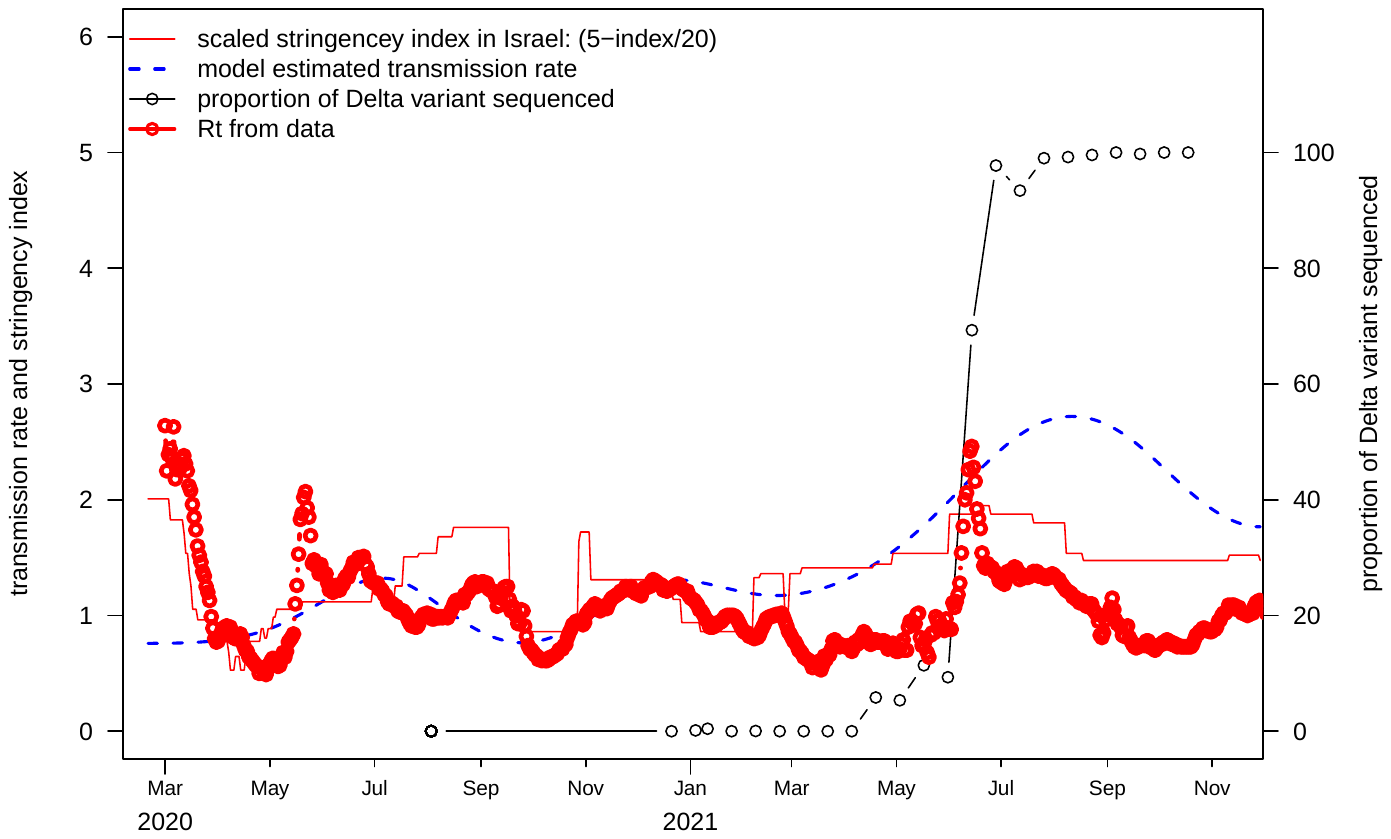

**Figure S6.** A comparison of the stringency index (100-index)/5 with the estimated transmission rate (in unit of $\beta/\gamma$), and proportion of Delta variant sequenced. The peak of the transmission rate in July-September 2021 is likely a combined effect of the relaxing of stringency and invasion of the more transmissible variant.

1.	Israel urges vaccination for all teens, citing Delta variant.Reuters website.

2.	Mallapaty S. Will COVID become a disease of the young? Nature. 2021;595(7867):343-4.

3.	What to Know About Breakthrough COVID-19 Cases Health Essentials: Health Essentials; 2021 [Available from: <https://health.clevelandclinic.org/breakthrough-covid-cases/>.

4.	Hale T, Angrist N, Goldszmidt R, Kira B, Petherick A, Phillips T, et al. A global panel database of pandemic policies (Oxford COVID-19 Government Response Tracker). Nature Human Behaviour. 2021;5(4):529-38.

5.	Toby Phillips HT. Methodology for Calculating Indices. 2021.

6.	Toby Phillips HT. Codebook for the Oxford Covid-19 Government Response Tracker. 2021.
